# Cheese consumption and risk of diabetic retinopathies: A Mendelian Randomization study

**DOI:** 10.1101/2023.08.18.23294288

**Authors:** Zenan Lin, Di Hu, Junhong Jiang, The μ-Biomedical Data Investigating Group (Mu-BioDig)

## Abstract

**Objectives:** Diabetic retinopathy (DR) is the leading cause of visual loss in working-age adults worldwide. Cheese is a widely consumed dairy product, and cheese intake has various health benefiting effects. This study aimed to use Mendelian randomization (MR) to investigate the impact of cheese consumption on DR.

**Methods:** The genome-wide association study (GWAS) summary statistic generated from 451486 European descent was introduced to identify the valid instrumental variables (IVs) for cheese intake. As the outcomes, the GWAS data of three outcome traits (DR, proliferative diabetic retinopathy or PDR, diabetic maculopathy or DMP) were obtained from the FinnGen research project. Multivariable MR (MVMR) analysis was also conducted to determine whether the causal associations were affected by the common risk factors of DR, such as Body Mass Index, systolic blood pressure (SBP), triglycerides (TG), HDL cholesterol and hemoglobin A1c (HbA1c) levels.

**Results:** The cheese consumption (per 1-SD increase) was found to be associated with a decreased risk of DR (OR=0.701, *P* =0.011), PDR (OR=0.671, *P*=0.020), and DMP (OR=0.357, *P*=0.001). The MVMR analyses demonstrated that the TG level did not affect the causal associations with the decreased risks of DR and PDR. The study on DMP proved that the causal relations were independent from the effect of SBP, TG, and HbA1c levels.

**Conclusions:** Genetic evidence suggested that cheese consumption was causally associated with the decreased risk of DR. Given widespread cheese consumption, this could have significant implications for global health burdens from DR.

## 1 Introduction

Diabetic retinopathy (DR), the leading cause of visual loss in working-age adults globally, is the most common vascular complication in diabetes mellitus.^1^ Proliferative diabetic retinopathy (PDR) and diabetic maculopathy (DMP) are the primarily vision-threatening stages of DR and lead to severe visual impairment, substantially diminishing vision-related quality of life.^2^ As the prevalence of diabetes grows yearly, the number of people with DR and the associated healthcare burden are expected to increase dramatically.^3^ Although DR can be treated by retinal laser photocoagulation, intravitreal injections, and vitrectomy; the efficacy of these strategies remains insufficient.^4, 5^ Therefore, to prevent the onset of DR as well as to delay the progression from early DR to vision-threatening DR are meaningful for alleviating the burden of DR-related blindness. Identifying causal and modifiable risk factors for DR is crucial for providing preventive strategies.

Dietary factors are the key modifiable risk factors that play essential roles in the preventing and managing diabetes.^6, 7^ However, compared with diabetes, little is known about the role of diet in the onset and development of DR.^8^ Cheese is a staple dairy product that has been reported to be associated with a lower risk of diabetes.^9^ Recently, a large observational study on diabetic patients found that higher consumption of cheese may reduce the risk of DR progression.^10^ Nevertheless, due to the potential confounding and reverse causation from observational studies,^11^ the causal relationship between cheese consumption and DR could not be concluded.

Mendelian randomization (MR) analysis is a powerful statistical method that exploits the random assortment of genetic variants during meiosis to assess the potential causal effect.^11, 12^ As an alternative to randomized clinical trials, MR analysis offers an opportunity to investigate the causal effect efficiently. It has become widely used to assess the causal associations of risk factors in diabetes.^13, 14^ To the best of our knowledge, so far no MR study has been conducted to investigate the causal role of cheese intake on DR.

## 2 Materials and methods

### 2.1 The genetic instrumental variables (IVs) for the consumption of cheese

In order to select the valid IVs for the consumption of cheese, the relevant genome-wide association studies (GWAS) summary dataset of the UK Biobank was acquired and analyzed via the IEU Open GWAS Project (https://gwas.mrcieu.ac.uk/datasets/, GWAS ID: ukb-b-1489).^15, 16^ The basic features of the dataset were described in Table 1 and Figure 1.

**Table 1.**
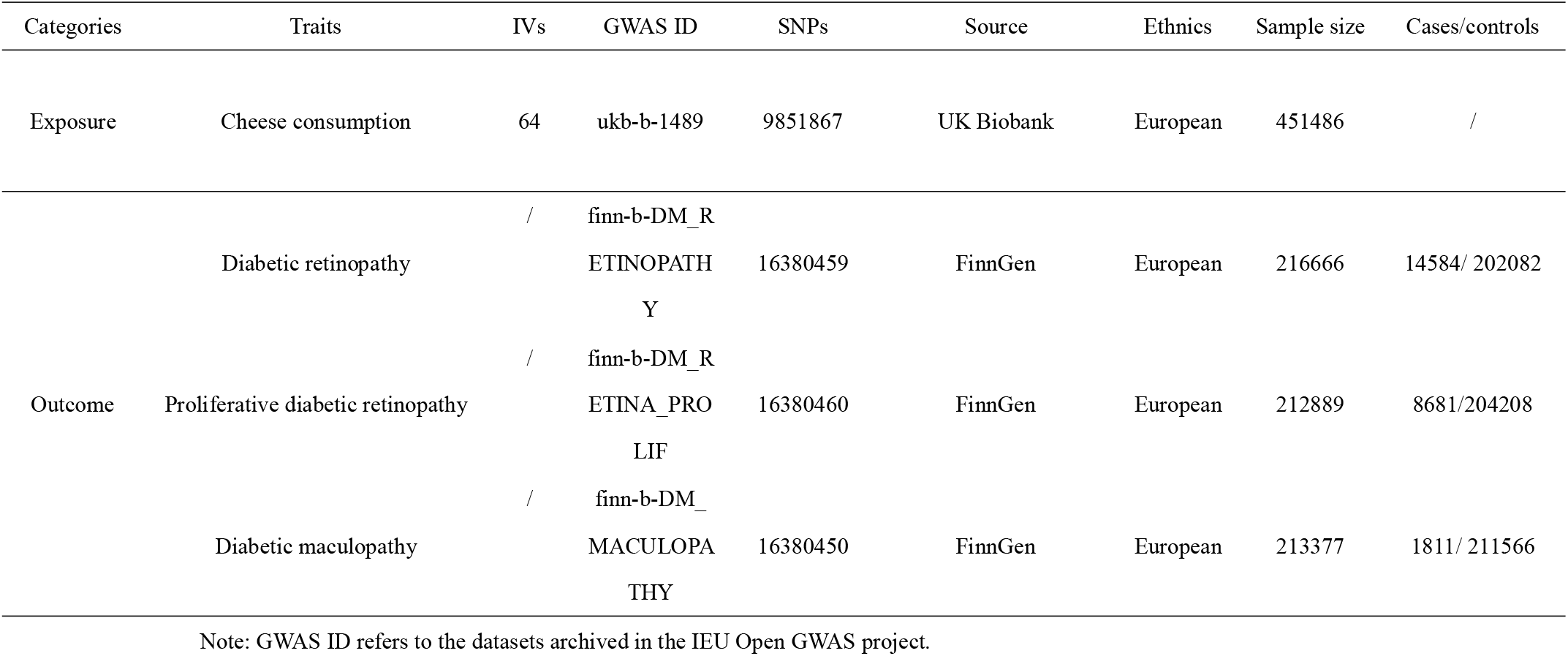
The basic information of the datasets of exposures and outcomes.

**Fig 1.**
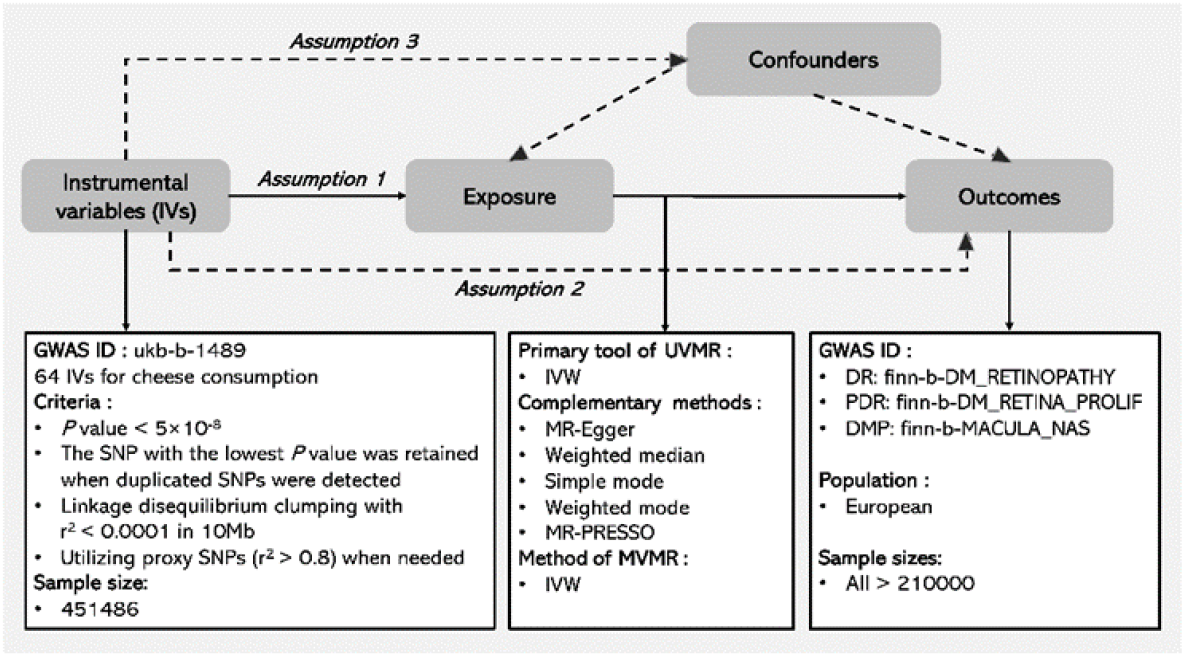
The workflow describing the MR analysis on the causal association between cheese consumption and DR risks.

According to the MR methodology, three key assumptions are: 1) the included IVs are robustly associated with the exposure trait; 2) they are associated with the outcome only through the exposure; 3) they are independent from the confounders. (See Figure 1.) In order to identify the independent SNPs, we selected the SNPs with *P* values less than 5×10^−8^ for the linkage disequilibrium clumping (r^2^ < 0.001 within 10000Kb, and using the 1000 Genomes Project Phase 3 (EUR) as the reference panel). When duplicated SNPs were detected, the one with the lowest *P* value was retained. When the SNPs were unavailable in the outcome’s dataset, the proxy SNPs (r^2^ > 0.8) were utilized for further analyses. Finally, 64 IVs for cheese consumption were identified to be valid IVs. (See Supplementary Table S1.) We calculated the F statistics to assess the association strength between each SNP and its corresponding trait. The results indicated sufficient statistical power for all SNPs as their F statistics (see Supplementary Table S1) were more significant than 10.^17^ The analyzing procedures were performed with the R (version 4.1.3) packages ieugwasr (version 0.1.5) and TwoSampleMR (version 0.5.6).^15^

### 2.2 The basic information of three outcome datasets

The GWAS datasets for diabetic retinopathy (DR), proliferative diabetic retinopathy (PDR) and diabetic maculopathy (DMP) were from the Finnish biobank project FinnGen (https://www.finngen.fi/fi). Their GWAS data was accessed via the platform of the IEU OpenGWAS project (https://gwas.mrcieu.ac.uk/datasets, GWAS ID: “finn-b-DM_RETINOPATHY”, “finn-b-DM_RETINA_PROLIF”, “finn-b-DM_MACULOPATHY”).^15, 16^ The characteristics of the datasets were shown in Table 1 and Figure 1.

### 2.3 Univariable and multivariable MR analysis

In the univariable MR (UVMR) analysis, the IVW approach was chosen as the major method, and other analytical tools such as the MR Egger, weighted median, weighted mode, simple mode, and MR-PRESSO were conducted to evaluate the robustness of the results. Moreover, the multivariable MR (MVMR) studies were introduced to explore the potential mechanism through which cheese consumption may influence the DR risk. The common risk factors for DR such as Body Mass Index (BMI), systolic blood pressure (SBP), triglyceride (TG), high-density lipoprotein (HDL) cholesterol, low-density lipoprotein (LDL) cholesterol, and hemoglobin A1c (HbA1c) levels were introduced into the MVMR analyses. The GWAS summary statistics of the common risk factors were acquired from the platform of the IEU OpenGWAS project (https://gwas.mrcieu.ac.uk/datasets, GWAS ID: “ukb-b-19953”, “ieu-b-38”, “ieu-b-111”, “ieu-b-109”, “ieu-b-110” and “ieu-b-103”, respectively).

### 2.4 Statistical analysis

Briefly, the IVW method was the major MR analytical tool, and complemented by other approaches such as the MR-Egger, simple mode, weighted median, weighted mode, and MR-PRESSO. The MVMR analysis was conducted to explore the potential mechanism through which the cheese intake may function. All analyses were conducted with R packages MRPRESSO (version 1.0), ieugwasr (version 0.1.5), and TwoSampleMR (version 0.5.6).^15, 16, 18^ A *P* value less than 0.05 was considered as statistically significant.

## 3 Results

### 3.1 The basic features of the included datasets for exposure and outcomes

Three GWAS datasets for the cheese consumption contained 9851867 SNPs’ data covering more than 42000 European participants. (See Table 1. and Figure 1.) As described in the method’s part, 64 IVs were regarded as valid IVs for cheese consumption. The data originated from the studies of the UK Biobank. Three DR-associated datasets from the FinnGen project were selected as the outcome cohorts for further analyses. The sample sizes for DR, PDR, and DMP were 216666 (14584 cases and 202082 controls), 212889 (8681 cases and 204208 controls), and 213377 (1811 cases and 211566 controls), respectively.

### 3.2 The UVMR analysis from cheese consumption to the risks of three diabetic retinopathies

The results of IVW analyses suggested that a 1-SD increase in cheese consumption was causally associated with a 29.9% decrease in the overall risk of DR (Odds ratio or OR=0.701, *P* =0.011), a 32.9% decrease in the PDR risk (OR=0.671, *P* =0.020) and a 64.3% decrease in the DMP risk (OR=0.357, *P* =0.001). Consistently, the sensitivity studies using MR Egger, Simple mode, Weighted median, Weighted mode, and MR-PRESSO also indicated cheese consumption to be a protective role for DR, PDR, and DMP, though some of them were not statistically significant. (See Table 2. and Figure 2.) While the *P* for Cochran’s Q in the IVW analysis on DR was less than 0.05, no heterogeneities were identified to exist in the studies on PDR (*P* for Cochran’s Q=0.078) and DMP (*P* for Cochran’s Q=0.588). (See Table 2. And Figure 2.) No obvious directional pleiotropies were detected in three exposure-outcome pairs since their Egger intercept terms were all centered at the origin (0.006 for DR, 0.001 for PDR, and -0.003 for DMP). Though the MR-PRESSO global studies also suggested no influence for the pleiotropy was observed in the analyses on PDR (*P*-global=0.052) and DMP (*P*-global=0.419), the investigation on DR implied a potential pleiotropy (*P*-global=0.038). The Leave-one-out tests demonstrated that no single SNP could greatly influence the overall estimates. (See Figure 2.)

**Table 2.**
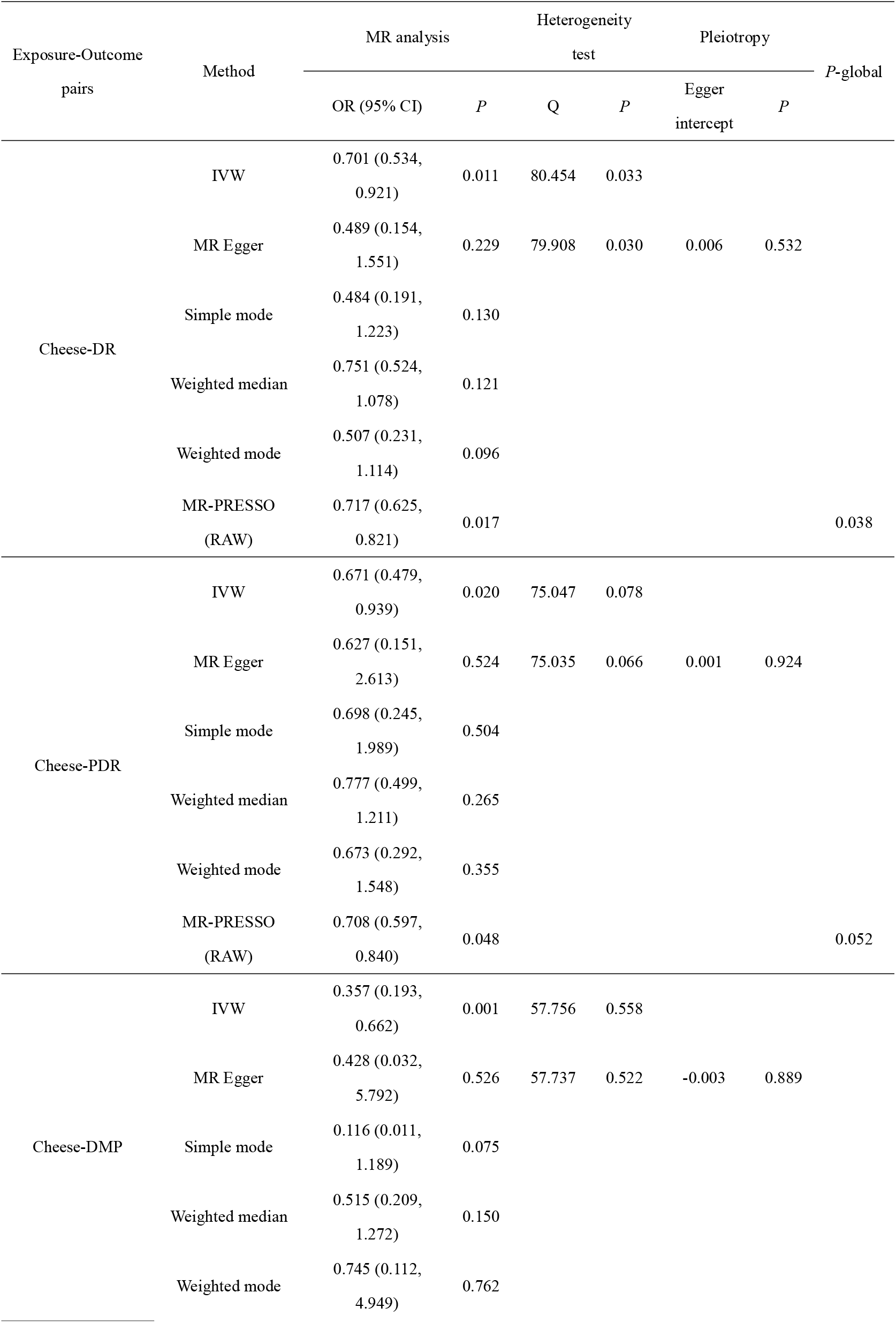
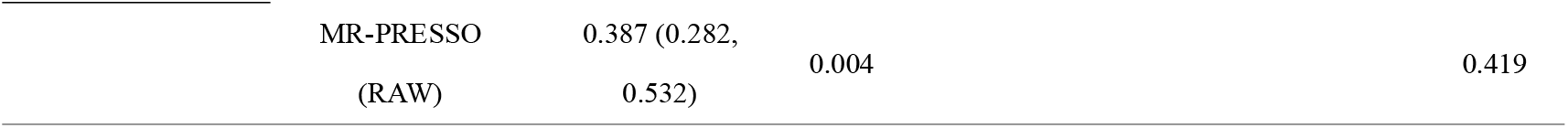
The MR analyses of the causal associations between the consumption of cheese and diabetic retinopathies.

**Fig 2.**
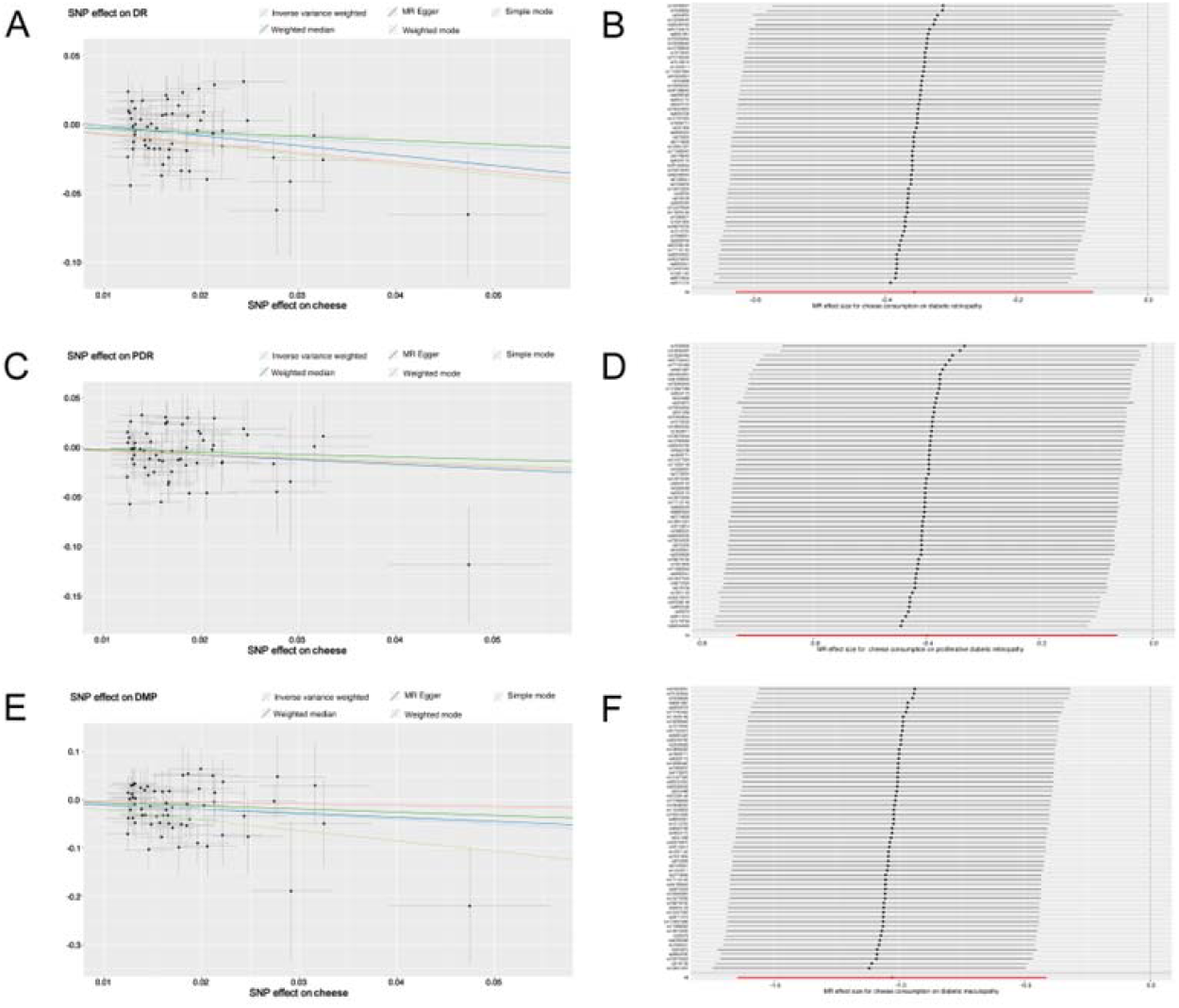
(A) Scatter plots of the genetic risks of the consumption of cheese on DR; (B) Forest plot of the Leave-one-out test on the MR study of DR; (C) Scatter plots of the genetic risks of the consumption of cheese on PDR; (D) Forest plot of the Leave-one-out test on the MR study of PDR; (E) Scatter plots of the genetic risks of the consumption of cheese on DMP; (F) Forest plot of the Leave-one-out test on the MR study of DMP.

### 3.3 The MVMR analysis from the cheese consumption to the risk of three diabetic retinopathies

As the UVMR suggested the potential causal associations between the consumption of cheese and the risk of three diabetic retinopathies, the MVMR was introduced to investigate the potential mechanisms through which the exposure may influence the outcomes.

We found that the TG level did not affect the causal associations with the risks of DR and PDR. (See Figure 3.) The study on DMP proved that the causal relationship was independent from the effect of SBP, TG, and HbA1c levels. (See Figure 3.)

**Fig 3.**
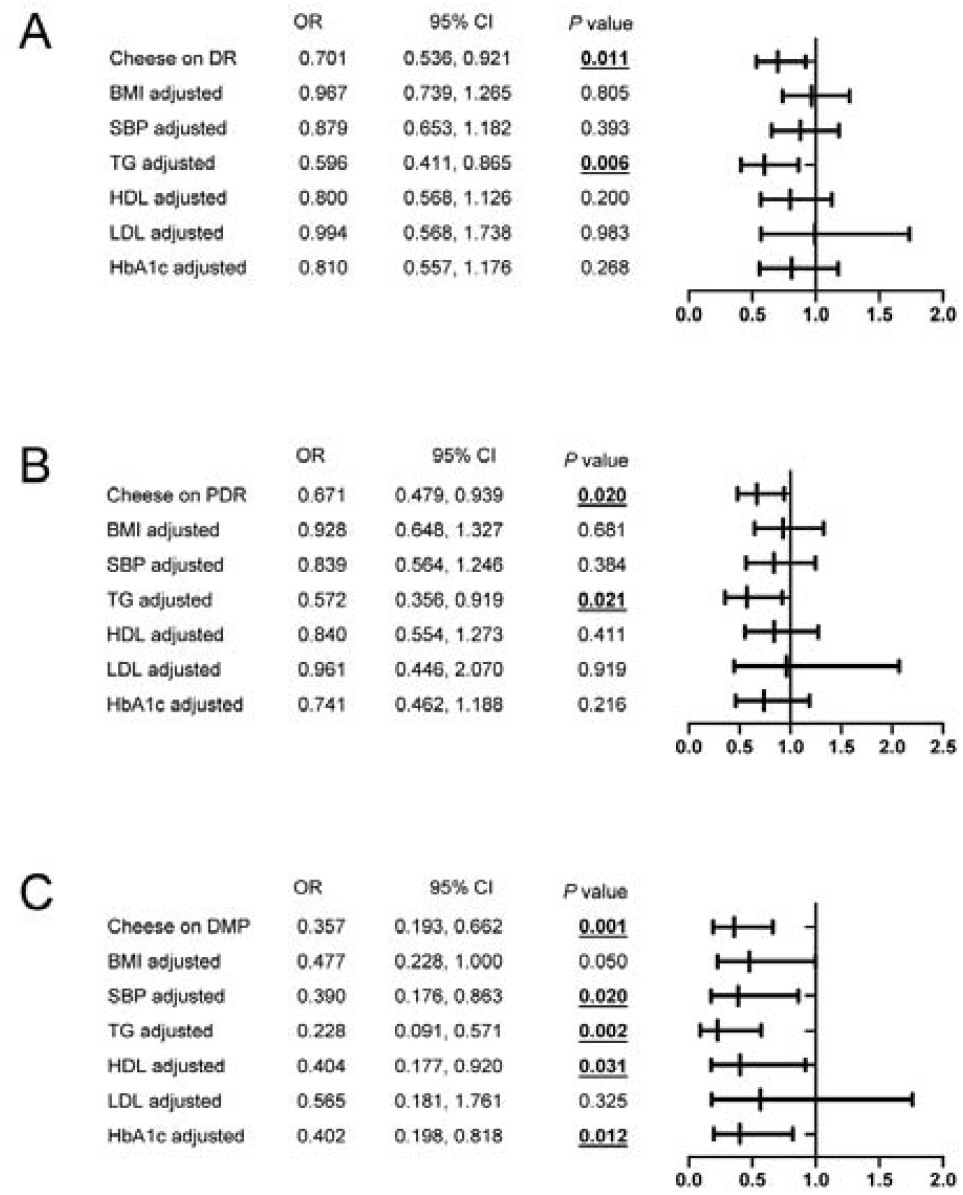
MVMR analyses of causal associations between cheese consumption and DR (A), PDR (B) and DMP (C) adjusted for common risk factors.

## 4 Discussion

In this two-sample MR study, we explored the causal effects of cheese consumption on diabetic retinopathies by leveraging large-scale GWAS data. This is the first study to show that consuming more habitual cheese was associated with a lower risk of developing diabetic retinopathies, providing a potential new treatment option for DR. Cheese is one of the most popular dairy products in the world and contains lots of nutrients. In recent years, numerous studies have been carried out to investigate the association between cheese consumption and diabetes risk. Their results revealed that eating cheese is inversely associated with diabetes.^19-22^ For instance, a large prospective case-cohort study across 8 European countries found a modest inverse association of cheese intake with diabetes.^20^ In a systemic review and meta-analysis on cohort studies, Aune and colleagues also identified the inverse association between cheese intake and the risk of type 2 diabetes.^21^ Interestingly, Von Post-Skagegard et al. reported that the protein in cheese has much stronger and quicker-acting glucoregulatory effects than fish protein and plant protein.^22^ Though previous works have implied the negative association between cheese consumption and diabetes risk; there has been little research exploring the relationship between eating cheese and DR risk. In this MR study, we confirmed that having cheese may be a protective factor for DR. This is in agreement with the results of prior studies on diabetes.

Cheese includes abundant calcium, vitamin D, whey protein, and magnesium. Multiple studies have demonstrated that all the substances as mentioned above are associated with reduced diabetes risk.^23-27^ This may partially explain the negative correlation between eating cheese and diabetes risk.

Despite the numerous studies on diabetes, to be best of our knowledge, only one published investigation has assessed the association between cheese intake and DR risk by observing the 10-year incidence of DR in diabetic patients in Australia.^28^ The study showed that cheese consumption could diminish the risk of DR in diabetic patients. This conclusion stands in line with our current work.

Higher BMI, SBP, HbA1c, LDL, and HDL cholesterol levels were acknowledged as risk factors for DR.^29^ Concurrently, these risk factors for DR are influenced by cheese intake: First, in systemic parameters, cheese intake was shown to reduce BMI in a randomized controlled trial.^30^ And a recent MR report demonstrated that cheese consumption was causally associated with a lower BMI.^31^ Furthermore, the inverse correlation between SBP and cheese intake has also been reported.^32^ Secondly, among circulating metabolites, HbA1c has been confirmed by a large RCT meta-analysis to be negatively associated with dairy product intake like cheese.^33^ Moreover, multiple RCTs showed that cheese intake could decrease the levels of LDL cholesterol concentration,^34, 35^ and increase the HDL-cholesterol concentration levels significantly.^36^ In concordance with those of previous studies, our MVMR results indicated that cheese consumption might function to protect DR by affecting these risk factors (see Figure 4). On the other hand, cheese consumption has been proven to be able to alleviate the oxidative stress^37^ and inflammatory response,^38^ which are the significant factors in the initiation and progression of DR.^39, 40^ This may also be one of the underlying mechanisms that cheese consumption suppresses DR (see Figure 4).

**Fig 4.**
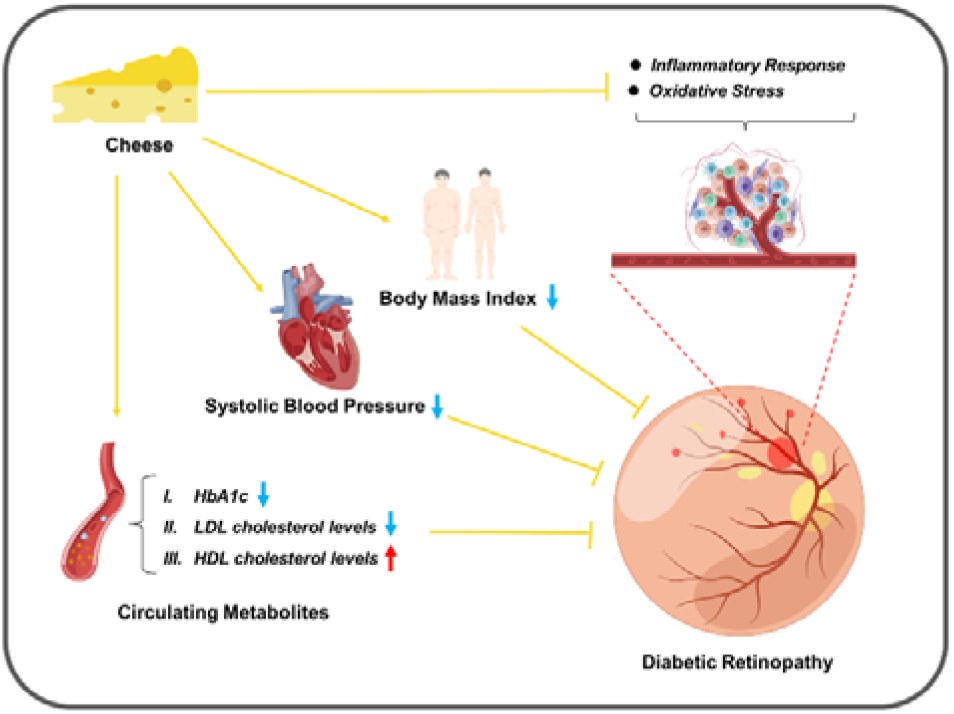
Diagram showing the possible mechanism by which cheese consumption reduces the risk of DR.

Though our work confirmed the causality between cheese consumption and DR risk through the MR framework, it is still limited by the following aspects. First, in the current work, cheese was investigated as a general subject instead of different cheese subtypes. The study with more accurate cheese subclassifications will offer a more precise and robust conclusion as different cheese subtypes vary in the health-associated nutrient components. Second, all the GWAS data analyzed originated from individuals of European descent which may impair the generalizability of the conclusion.

In general, our study firstly reported the inverse association between eating cheese and DR risk from a MR perspective. This finding implies that daily cheese intake may be regarded as a modifiable approach to decrease the DR risk.

## Supporting information

Table S1. 64 IVs for cheese consumption

## Data Availability

All data produced in the present study are available upon reasonable request to the authors

## Conflict of Interest

The authors declare no conflict of interest.

## Acknowledgement

We want to thank to the UK biobank and FinnGen project as well as the participants who generously made the GWAS summary statistics of cheese intake and diabetic retinopathies publicly available. No fund was obtained for this work.

## Ethics Approval

The used GWAS data were publicly available and approved by their corresponding institutions. A local ethics approval for the current work is not required.

## Data availability

The GWAS data were publicly available and their sources were described appropriately in the manuscript.

## μ-Biomedical Data Investigation Group

Zenan Lin^1^, Junhong Jiang^1^, Di Hu^2^, and Qi Zhang^3^

^1^The Department of Ophthalmology, Shanghai General Hospital, Shanghai Jiao Tong University School of Medicine, Shanghai, 200080, China.

^2^The Department of Ophthalmology, Children’s Hospital of Fudan University, No.399 Wanyuan Road, Shanghai, 201102, China.

^3^The Department of Neurology, The Affiliated Taizhou People’s Hospital of Nanjing Medical University, Taizhou School of Clinical Medicine, Nanjing Medical University, Taizhou, Jiangsu, 225300, China.

## Notes

### Competing Interest Statement

The authors have declared no competing interest.

### Funding Statement

This study did not receive any funding

### Author Declarations

IEU Open GWAS Project (https://gwas.mrcieu.ac.uk/datasets/

## References

1. Fong DS, Aiello L, Gardner TW, et al. Diabetic retinopathy. Diabetes care 2003;26 Suppl 1:S99–s102.

2. Klein R, Klein BE, Moss SE. Visual impairment in diabetes. Ophthalmology 1984;91:1–9.

3. Causes of blindness and vision impairment in 2020 and trends over 30 years, and prevalence of avoidable blindness in relation to VISION 2020: the Right to Sight: an analysis for the Global Burden of Disease Study. The Lancet Global health 2021;9:e144–e160.

4. Mohamed Q, Gillies MC, Wong TY. Management of diabetic retinopathy: a systematic review. Jama 2007;298:902–16.

5. Elman MJ, Qin H, Aiello LP, et al. Intravitreal ranibizumab for diabetic macular edema with prompt versus deferred laser treatment: three-year randomized trial results. Ophthalmology 2012;119:2312–8.

6. Ojo O. Dietary Intake and Type 2 Diabetes. Nutrients 2019;11.

7. Ley SH, Hamdy O, Mohan V, Hu FB. Prevention and management of type 2 diabetes: dietary components and nutritional strategies. Lancet (London, England) 2014;383:1999–2007.

8. Dow C, Mancini F, Rajaobelina K, et al. Diet and risk of diabetic retinopathy: a systematic review. 2018;33:141–156.

9. Díaz-López A, Babio N, Martínez-González MA, et al. Mediterranean Diet, Retinopathy, Nephropathy, and Microvascular Diabetes Complications: A Post Hoc Analysis of a Randomized Trial. Diabetes care 2015;38:2134–41.

10. Yan X, Han X, Wu C, et al. Does daily dietary intake affect diabetic retinopathy progression? 10-year results from the 45 and Up Study. 2020;104:1774–1780.

11. Davey Smith G, Hemani G. Mendelian randomization: genetic anchors for causal inference in epidemiological studies. Human molecular genetics 2014;23:R89–98.

12. Cui Y, Si W, Zhu C. Alcohol Consumption and Mild Cognitive Impairment: A Mendelian Randomization Study from Rural China. 2022;14.

13. Pedron S, Kurz CF, Schwettmann L, Laxy M. The Effect of BMI and Type 2 Diabetes on Socioeconomic Status: A Two-Sample Multivariable Mendelian Randomization Study. 2021;44:850–852.

14. Manousaki D, Harroud A. Vitamin D levels and risk of type 1 diabetes: A Mendelian randomization study. 2021;18:e1003536.

15. Hemani G, Zheng J, Elsworth B, et al. The MR-Base platform supports systematic causal inference across the human phenome. Elife 2018;7.

16. Elsworth B, Lyon M, Alexander T, et al. The MRC IEU OpenGWAS data infrastructure. bioRxiv 2020:2020.08.10.244293.

17. Burgess S, Butterworth A, Thompson SG. Mendelian randomization analysis with multiple genetic variants using summarized data. Genet Epidemiol 2013;37:658–65.

18. Verbanck M, Chen C-Y, Neale B, Do R. Detection of widespread horizontal pleiotropy in causal relationships inferred from Mendelian randomization between complex traits and diseases. Nature Genetics 2018;50:693–698.

19. Poppitt SD. Cow’s Milk and Dairy Consumption: Is There Now Consensus for Cardiometabolic Health? Front Nutr 2020;7:574725.

20. Sluijs I, Forouhi NG, Beulens JW, et al. The amount and type of dairy product intake and incident type 2 diabetes: results from the EPIC-InterAct Study. Am J Clin Nutr 2012;96:382–90.

21. Aune D, Norat T, Romundstad P, Vatten LJ. Dairy products and the risk of type 2 diabetes: a systematic review and dose-response meta-analysis of cohort studies. Am J Clin Nutr 2013;98:1066–83.

22. von Post-Skagegård M, Vessby B, Karlström B. Glucose and insulin responses in healthy women after intake of composite meals containing cod-, milk-, and soy protein. European Journal of Clinical Nutrition 2006;60:949–954.

23. Pittas AG, Lau J, Hu FB, Dawson-Hughes B. The role of vitamin D and calcium in type 2 diabetes. A systematic review and meta-analysis. J Clin Endocrinol Metab 2007;92:2017–29.

24. Mitri J, Muraru MD, Pittas AG. Vitamin D and type 2 diabetes: a systematic review. Eur J Clin Nutr 2011;65:1005–15.

25. Dong JY, Xun P, He K, Qin LQ. Magnesium intake and risk of type 2 diabetes: meta-analysis of prospective cohort studies. Diabetes Care 2011;34:2116–22.

26. Volpe SL. Magnesium, the metabolic syndrome, insulin resistance, and type 2 diabetes mellitus. Crit Rev Food Sci Nutr 2008;48:293–300.

27. Mozaffarian D, Cao H, King IB, et al. Trans-palmitoleic acid, metabolic risk factors, and new-onset diabetes in U.S. adults: a cohort study. Ann Intern Med 2010;153:790–9.

28. Yan X, Han X, Wu C, et al. Does daily dietary intake affect diabetic retinopathy progression? 10-year results from the 45 and Up Study. Br J Ophthalmol 2020;104:1774–1780.

29. Ting DS, Cheung GC, Wong TY. Diabetic retinopathy: global prevalence, major risk factors, screening practices and public health challenges: a review. Clin Exp Ophthalmol 2016;44:260–77.

30. Sharafedtinov KK, Plotnikova OA, Alexeeva RI, et al. Hypocaloric diet supplemented with probiotic cheese improves body mass index and blood pressure indices of obese hypertensive patients--a randomized double-blind placebo-controlled pilot study. Nutrition journal 2013;12:138.

31. Hu MJ, Tan JS, Gao XJ, Yang JG, Yang YJ. Effect of Cheese Intake on Cardiovascular Diseases and Cardiovascular Biomarkers. Nutrients 2022;14.

32. Schmidt KA, Cromer G, Burhans MS, et al. Impact of low-fat and full-fat dairy foods on fasting lipid profile and blood pressure: exploratory endpoints of a randomized controlled trial. 2021;114:882–892.

33. O’Connor S, Turcotte AF, Gagnon C, Rudkowska I. Increased Dairy Product Intake Modifies Plasma Glucose Concentrations and Glycated Hemoglobin: A Systematic Review and Meta-Analysis of Randomized Controlled Trials. Advances in nutrition (Bethesda, Md) 2019;10:262–279.

34. Hjerpsted J, Leedo E, Tholstrup T. Cheese intake in large amounts lowers LDL-cholesterol concentrations compared with butter intake of equal fat content. The American journal of clinical nutrition 2011;94:1479–84.

35. Brassard D, Tessier-Grenier M, Allaire J, et al. Comparison of the impact of SFAs from cheese and butter on cardiometabolic risk factors: a randomized controlled trial. The American journal of clinical nutrition 2017;105:800–809.

36. Thorning TK, Raziani F, Bendsen NT, Astrup A. Diets with high-fat cheese, high-fat meat, or carbohydrate on cardiovascular risk markers in overweight postmenopausal women: a randomized crossover trial. 2015;102:573–81.

37. Vasconcelos FM, Silva HLA, Poso SMV, et al. Probiotic Prato cheese attenuates cigarette smoke-induced injuries in mice. Food research international (Ottawa, Ont) 2019;123:697–703.

38. Tognocchi M, Conte M. Supplementation of Enriched Polyunsaturated Fatty Acids and CLA Cheese on High Fat Diet: Effects on Lipid Metabolism and Fat Profile. 2022;11.

39. Stitt AW, Curtis TM, Chen M, et al. The progress in understanding and treatment of diabetic retinopathy. Progress in retinal and eye research 2016;51:156–86.

40. Bryl A, Falkowski M, Zorena K. The Role of Resveratrol in Eye Diseases-A Review of the Literature. 2022;14.

